# Lipoprotein retention and inflammation due to regurgitant blood flow as part of the natural history of degenerative ascending aortic aneurysms

**DOI:** 10.1101/2024.07.29.24311194

**Authors:** David Freiholtz, Karin Lång, Otto Bergman, Christian Olsson, Malin Granbom Koski, Michael Dismorr, Cecilia Österholm, Kenneth Caidahl, Anders Franco-Cereceda, Per Eriksson, Anton Gisterå, Hanna M Björck

## Abstract

**BACKGROUND:** An abnormal accumulation of immune cells and a disrupted lipoprotein metabolism has previously been described as part of the pathogenesis of ascending aortic aneurysm in patients with tricuspid aortic valves. The factor driving the accumulation of immune cells remains unclear; however, it may be considered in light of the observation that proximal aortic dilatation often occurs alongside aortic regurgitation but rarely with aortic stenosis. In the present study we aim to investigate the natural history of ascending aortic aneurysm in patients with tricuspid aortic valves by assessing the association between aortic regurgitation and vascular deterioration.

**MATERIAL AND METHODS:** Patients tricuspid aortic valves undergoing elective open-heart surgery for ascending aortic- and/or aortic valve replacement were included. Aortic specimens from organ donors were obtained through the University of Miami Tissue Bank, USA. Protein expression/localization and differences in aortic intima-media gene expression were assessed using immunohistochemistry and transcriptomics, respectively. Ten-year aortic growth was measured using echocardiography. In total 142 patients were included across experiments (mRNA expression n=44, immunohistochemistry n=49, 10-year follow-up n=49).

**RESULTS:** Aortic regurgitation was associated with the presence of oxidized apolipoprotein B-containing lipoproteins and infiltrating CD68+ cells in the non-dilated ascending aortic media, which was not observed in aortas of patients with aortic stenosis. Assessing factors influencing lipoprotein retention showed increased levels of genes encoding core proteins of proteoglycans (*HSPG2, CSPG4, ACAN*, and *BGN*) in patients with regurgitant valves, compared with aortas from patients with stenotic valves. Moreover, dilated aortas of patients with aortic regurgitation exhibited higher levels of the receptor for oxidized low-density lipoprotein, *OLR1*, which correlated positively with inflammatory markers in both dilated and non-dilated aortas. Surgical replacement of regurgitant aortic valves mitigated long-term aortic growth, in contrast to replacement of stenotic valves, which was associated with continuous aortic dilation.

**CONCLUSIONS:** The natural history of ascending aortic aneurysm in patients with tricuspid aortic valves involves medial lipoprotein retention and oxidation with subsequent *OLR1*-driven pathological inflammation, and can be mitigated by replacement of the regurgitant aortic valve.

## Introduction

Degenerative ascending aortic aneurysm is a silent, potentially fatal disease^1^. It manifests as a progressive enlargement of the aortic lumen due to destructive changes in the aortic connective tissue. Pathologically it is characterized by vascular smooth muscle cell loss, extracellular matrix remodeling and low-grade inflammation^2–4^. The mechanism underlying its onset remains elusive, and no medical therapy or prognostic factor has yet been identified. Interestingly, non-canonical atherosclerotic processes have recently been described in the aneurysmal intima/media of patients with tricuspid aortic valves, signified by a pathological accumulation of macrophages and a disturbed lipid metabolism^5^. In our recent work, we could further enhance these findings by the identification infiltrating CD68+ immune cells expressing Secreted Phosphoprotein 1 that correlated with aortic dimensions and an augmented expression of inflammatory markers^6^. The mechanism enabling immune cells to migrate into to the sub-intimal space to facilitate inflammation initiation remains, however, unknown.

Notably, patients with tricuspid aortic valves undergoing surgery for ascending aortic aneurysm often present with concomitant aortic valve regurgitation^7,8^. Equivalent patients with aortic stenosis, on the other hand, seem to be protected against proximal aneurysm development^9^. Regurgitation involves an inadequate closure of the aortic valve, causing backflow and flow-reversals in the proximal aorta^10^ with less laminar flow and decreased anti-inflammatory endothelial signaling^11^. This obviously alters endothelial cell function and facilitates infiltration of *e.g*., lipoprotein and leukocytes into the aortic wall^12^. Moreover, mild medial degeneration and smooth muscle cell phenotypic alterations^13^ has previously been reported in patients with aortic regurgitation without ascending aortic dilatation^14–16^, and aortic regurgitation *per se* has been associated with more adverse outcomes following ascending aortic surgery^8,17^. In the present study, we sought to investigate the natural history of ascending aortic aneurysm formation in patients with tricuspid aortic valves, hypothesizing that regurgitant blood flow in the proximal aorta is associated with lipoprotein infiltration and retention, and consequently, activation of inflammation and medial remodeling.

## Material and Methods

### Patients

The present study includes patients from the Advanced Study of Aortic Pathology (ASAP) and Disease of the Aortic Valve, Ascending Aorta and Coronary Artery (DAVAACA) cohorts^18,19^. In short, all patients underwent elective open-heart surgery for aortic valve disease and/or ascending aortic aneurysm at the Karolinska University Hospital, Stockholm, Sweden. Tissue biopsies were collected from the anterior part of the dilated ascending aorta, or at the site of aortotomy a few centimeters above the aortic valve from patients undergoing isolated aortic valve replacement. Biopsies of the internal thoracic artery were collected from patients included in the ASAP cohort. Ascending aortas with diameters > 45 mm were classified as dilated. Ascending aortic diameters were determined by perioperative transesophageal echocardiography. Patients with syndromic aortic pathologies or ascending aortic dissection were excluded. Analogous control tissues were acquired from organ donors with no documented history of valvular heart disease, through collaboration with Nova Southeastern University and the University of Miami Tissue Bank. A description of the control cohort can be found elsewhere^20^. Ethical approval for the use of all tissues included in the study were obtained from the regional ethics review committee in Stockholm. Non-overlapping patients were selected for mRNA expression (n=44) and immunohistochemistry analysis, respectively (in total n=49), supplementary table S1.

### 10-year follow up

Patients included in the ASAP cohort were re-called 10 years after surgery and aortic diameters were measured using echocardiography in those accepting to participate in the follow-up. Of the initial 600 ASAP patients, 419 were alive at the time of the follow-up, of which 81.6% (n= 342) accepted to participate. Of these, 210 patients had undergone isolated valve replacement, 80 of which had a tricuspid aortic valve (remaining 130 patients had a bicuspid aortic valve and were not included). After exclusion of patients who have had an event in their ascending aorta prior to follow-up (n=4), or patients with incomplete echocardiographic measurements (n=9 at follow-up, n=18 pre-operatively)), a total of 49 patients remained (n=12 regurgitation and n=37 with stenosis) in the final analysis. Mean follow-up time from surgery to echocardiography was 10.3 years.

### mRNA extraction and gene expression analysis

The intima-media layer was separated from the aortic tissue through adventectomy and mRNA was extracted from the intima-media portion using the RNeasy kit (Qiagen). Global gene expression was measured using the Affymetrix GeneChip® Human Exon 1.0 ST array and protocols, as previously described^21^, in 23 patients with non-dilated aortas (n=11 with stenosis, and n=12 with regurgitation) and 21 patients with dilated aortas (n=20 with complete aortic diameters).

### Immunohistochemistry

Cryosections (8 µm) of aortic surgical biopsies were fixed in acetone and blocked with 10% goat serum with 0.1% Tween for 30 min. Sections were then incubated overnight at 4°C with primary antibodies: anti-apoB (1:1000), CD68 (1:100), OxLDL (1:500), and OLR1 (1:500). For ApoB, slides were incubated with biotinylated goat anti-mouse secondary antibody, followed by streptavidin 568-conjugated fluorophore. Autofluorescence was quenched with 0.01% Sudan black. OxLDL and LOX-1 were visualized with a 647 fluorophore-conjugated secondary anti-rabbit antibody, and CD68 with a 568 fluorophore-conjugated anti-mouse antibody. Nuclei were counterstained with 4′,6-diamidino-2-phenylindole (DAPI) and sections mounted in Fluoromount-G. Stainings were visualized using a laser scanning confocal microscope. Acetone-fixed cryosections were also stained with Oil-red O, counterstained with Hematoxylin-Q, and examined with a brightfield scanner.

A portion of the biopsies were fixed in 4% paraformaldehyde, embedded in paraffin, and sectioned at 5 µm. Immunostaining for CD68 was performed overnight at 4°C on deparaffinized sections after antigen retrieval, endogenous peroxidase quenching, and blocking with 20% goat serum. Biotinylated goat anti-mouse IgG was used as the secondary antibody (1:1,500) with avidin-biotin peroxidase complex added for visualization with DAB. Sections were counterstained with hematoxylin, rehydrated and mounted. Hematoxylin-eosin staining was performed following manufacturer instructions. Sections were examined using a brightfield scanner.

### Image analysis

Quantifications of signal intensity was performed in Fiji (Version 2.9.0, ImageJ2) upon sectioning subintimal and medial region for ApoB, oxLDL and LOX-1. CD68+ positive cell-ratio quantification was performed in Qupath (version 0.5.0, QuPath) averaging the ratio of positive cells to all cells in three randomly selected intima-media regions.

Hematoxylin-eosin was used to assess overall medial degeneration. Parameter definitions given by the consensus statement on noninflammatory aortic disease were adapted^22^. Briefly, the severity of medial disorganization was assessed by consecutive number of disrupted lamellar units in digital images. Severity was rated on a scale of 0 to 3, where 0 equals no apparent pathology, 1 equals 2-4 consecutive lamellar units affected, 2 corresponds to 5-9 and 3 represents >10.

### Statistical analysis

Baseline characteristics are presented as means (standard deviation) for continuous variables, and ratio (percentage) for categorical variables, with differences analyzed using Mann-Whitney U test and Fisher’s exact test for continuous and categorical variables, respectively. Differences in fluorescence intensity were compared using the Kruskal-Wallis test followed by Dunn’s multiple comparisons test, or using Brown-Forsythe and Welch ANOVA with Dunnett’s T3 multiple comparisons test. The Mann-Whitney U test was employed when appropriate. 10-year aortic growth was evaluated using Wilcoxon matched pairs signed rank test. Differential gene expression was analyzed using student’s t-test, assuming unequal variance. Functional enrichment was determined using the msigdbr package in Rstudio (v1.4). The Reactome canonical pathway was utilized to minimize redundancy. Correlations were assessed using Pearson’s correlation. A p-value <.05 was considered for statistical significance unless otherwise stated. Statistical analysis was carried out using GraphPad Prism 10 (v10.1.1) or Rstudio (v1.4).

## Results

### Aortic regurgitation is associated with infiltration of lipoproteins and immune cells in the non-dilated aortic wall

Immunohistochemical evaluation of aortic biopsies revealed striking differences in sub- intimal expression of apoB, Oil Red O+ lipids, oxLDL, and CD68+ macrophages (Fig. 1A) between patients with aortic regurgitation and those with aortic stenosis. Specifically, a gradually reduced staining intensity, 50-100 µm into the medial layer, of apoB lipoproteins, Oil Red O+ lipids, oxLDL, and CD68+ macrophages was exclusively seen in non-dilated aortas of patients with aortic regurgitation. The gradually reduced staining suggesting infiltration from the aortic lumen. We could not see any intimal or medial staining of any of the above-mentioned proteins in patients with aortic stenosis. To investigate whether the observed lipoprotein retention was restricted to the ascending aorta, we also examined biopsies obtained from the internal thoracic artery, showing no medial infiltration of apoB in either patients with stenosis or regurgitation (Fig. 1B). Notably, although not statistically significant, patients with stenotic valves showed a tendency towards higher plasma cholesterol levels (P=.06) and were older (P=.021) that patients with aortic regurgitation. Low-density lipoprotein cholesterol concentrations did not differ between groups (P=.74) (Table 1). Aortic biopsies obtained from organ donors without any documented valvulopathy showed sub-intimal staining of apoB but had no detectable levels of lipids or oxidized lipoproteins, indicating a lack of LDL-particle retention.

**Figure 1.**
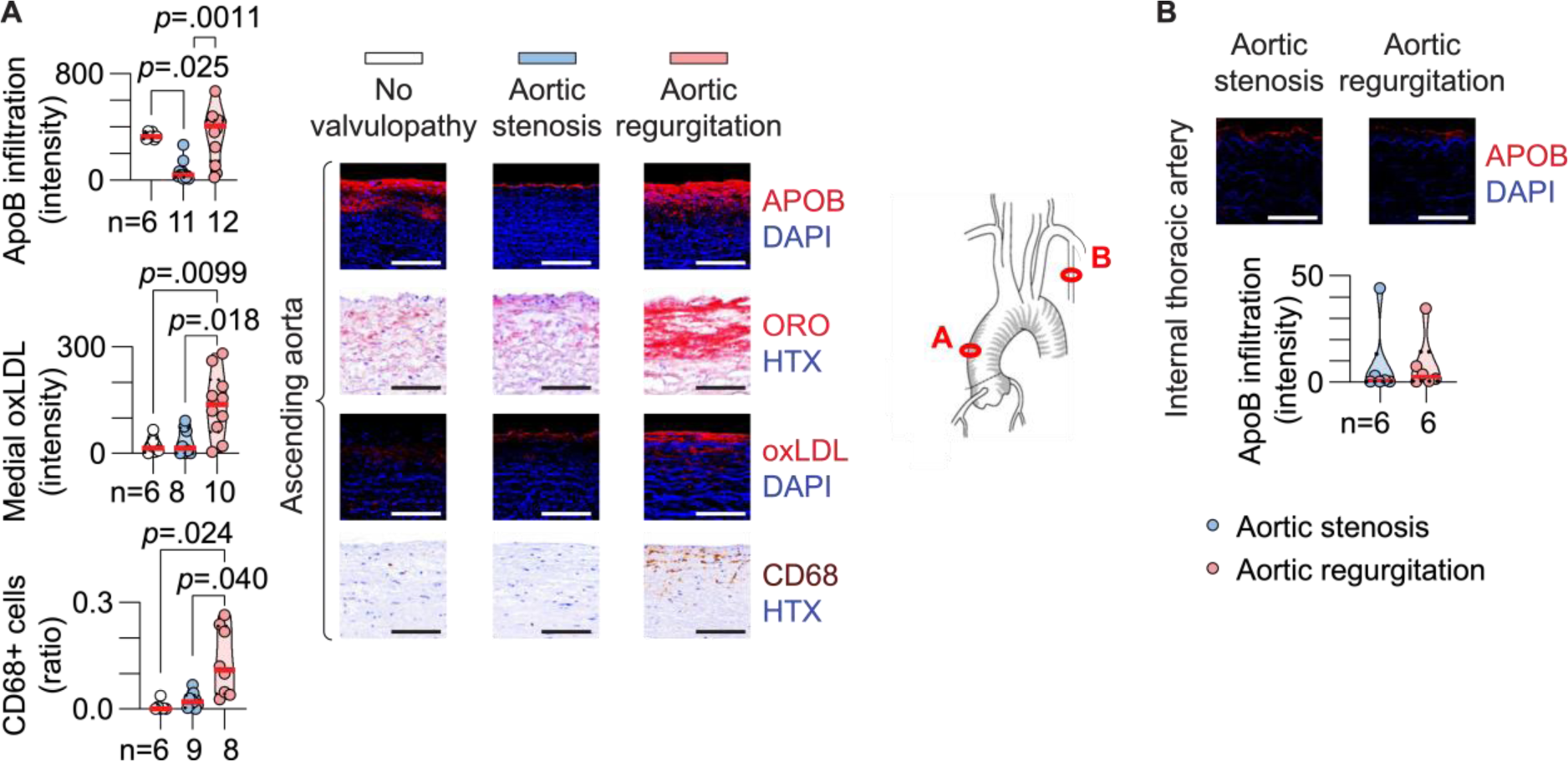
Infiltration of lipoproteins and inflammatory cells in the ascending aorta. (A) Immunofluorescent staining of apolipoprotein(apo)B, oxidized(ox)LDL, and immunohistological staining of CD68 in ascending aortic biopsies, and representative micrographs of Oil red O (ORO) staining. Kruskal-Wallis test with Dunn’s multiple comparisons test for apoB and Brown-Forsythe and Welch ANOVA with Dunnet’s T3 multiple comparisons test for oxLDL and CD68. **(B)** ApoB staining in internal thoracic arteries with quantification of staining intensity in media. Mann-Whitney test (*p*-value=.48). Scale bar 100 µm in all micrographs. DAPI=4′,6-diamidino-2-phenylindole, HTX=hematoxylin.

**Table 1.** Patient characteristics of individuals examined for ApoB-infiltration (immunohistochemistry).

|  | Aortic Stenosis | Aortic Regurgitation | P-value |
| --- | --- | --- | --- |
| **N** | 11 | 12 | - |
| **Age, years** | 65.08 (14.9) | 57.9 (11.3) | .021 |
| **Male sex** | 8 (73) | 8 (67) | 1.0 |
| **BSA, m<sup>2</sup>** | 1.98 (0.2) | 1.94 (0.2) | .52 |
| **Cholesterol, mmol/L** | 4.91 (1.3) | 4.16 (1.1) | .060 |
| **LDL-cholesterol, mmol/L** | 3.02 (1.2) | 2.3 (1) | .74 |
| **hsCRP, mg/L** | 2.4 (2.8) | 6.4 (16.2) | .37 |
| **Diabetes** | 0 (0) | 0 (0) | 1.0 |
| **Hypertension** | 6 (55) | 8 (67) | .68 |
| **Chronic inflammation** | 0 (0) | 0 (0) | 1.0 |
| **Vascular disease** | 4 (36) | 1 (8) | .16 |
| **Current smoker** | 6 (55) | 4 (33) | .41 |

Continuous variables are presented as mean (SD); ordinal variables are presented as n (ratio). Mann-Whitney and Fisher's exact test for numerical and categorical variables respectively. Body surface area, BSA; high-sensitivity C-reactive protein, hsCRP; low-density lipoprotein, LDL.

### Matrix remodeling may impact apoB lipoprotein retention in aortas of aortic regurgitation patients

In line with previous publications^11–13^, an increased medial degradation of the non-dilated ascending aorta, including fragmented elastic lamellae, was seen in patients with aortic regurgitation compared with patients with aortic stenosis (P=.039) (Fig. 2A). No medial degeneration was seen in organ donors devoid of documented valve disease (Fig. 2A). Global gene expression analysis revealed that genes associated with extracellular matrix (ECM) organization and ECM proteoglycans were significantly up-regulated in non-dilated aortas of patients with aortic regurgitation compared with aortic stenosis patients (Fig. 2B, top 100 differentially expressed genes are shown in supplementary table S2). Specifically, we observed increased mRNA levels of key genes encoding core proteins of LDL-binding proteoglycans^23^ (*HSPG2, CSPG4, ACAN*, and *BGN)* (Fig 2C), suggesting a milieu favoring lipoprotein retention.

**Figure 2.**
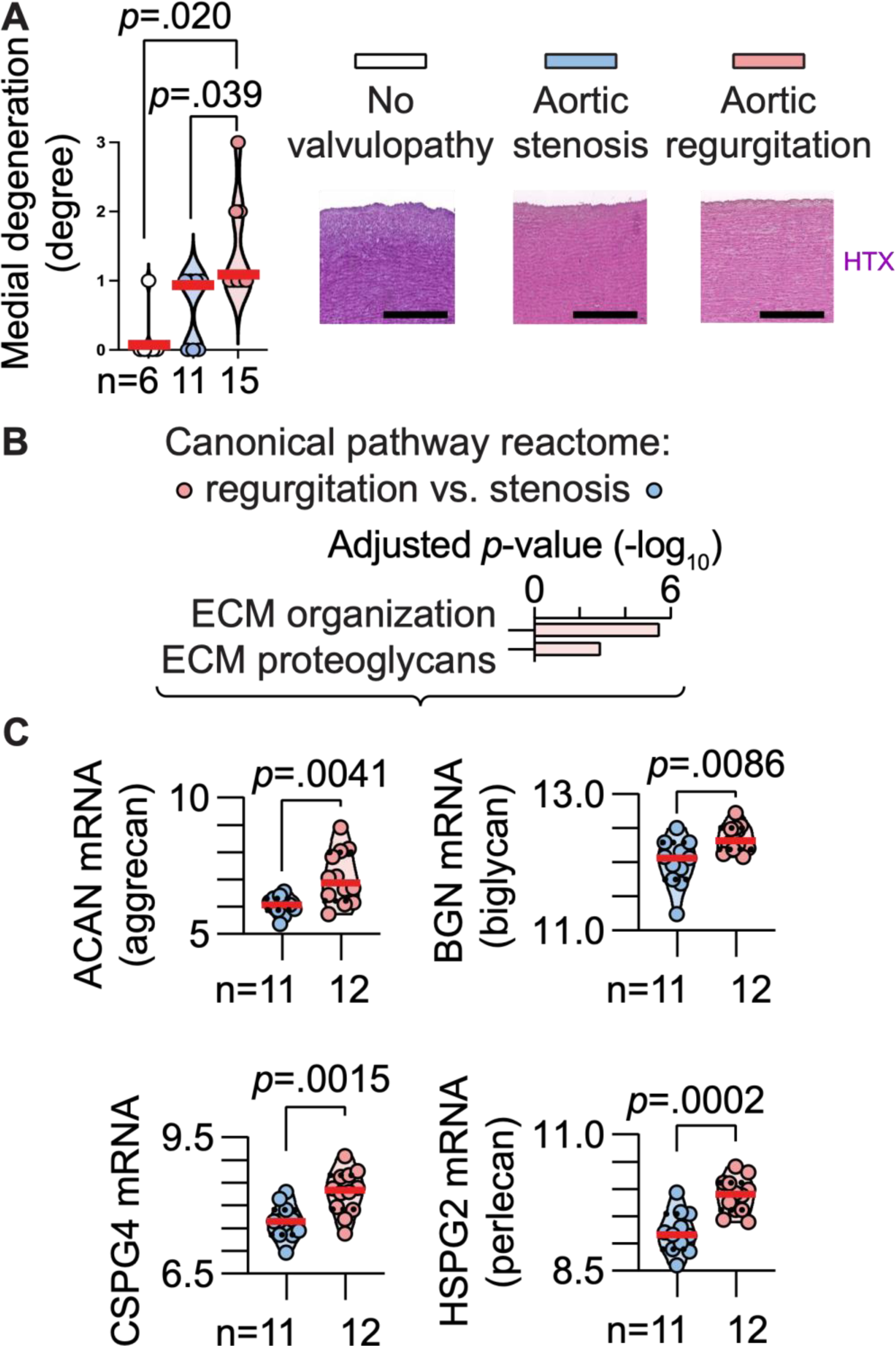
Matrix remodeling in the non-dilated ascending aorta associates with aortic regurgitation **(A)** Hematoxylin-eosin (HTX) staining of ascending aortic biopsies. Mann-Whitney test. Scale bar 250 µm in all micrographs. **(B)** Pathway analysis of differentially expressed genes between patients with aortic regurgitation vs. aortic stenosis, non-dilated aortas. **(C)** Proteoglycan mRNA levels in non-dilated aortic biopsies from patients with aortic regurgitation vs. stenosis. Unpaired t-test with Welch’s correction for mRNA level comparisons. ECM=extracellular matrix.

### Oxidized lipoprotein receptor 1 (OLR1) as a driver of degenerative ascending aortic aneurysm formation

To further clarify the role of lipoprotein oxidization in aneurysm formation, we conducted a functional enrichment analysis of differentially expressed genes between non-dilated and dilated ascending aortas of patients with aortic regurgitation. This showed an up-regulation of genes related to the innate immune system as key in the aneurysmal process. The most highly up-regulated genes were *CLEC5A, OLR1, TREM1, PTPRC*, and *CCR2* (Fig. 3A) (full list of genes within this set in supplementary table S3). Interestingly, GeneMania analysis showed that oxidized lipoprotein receptor 1 (*OLR1*) physically interacts with apoB (Fig. 3B). OLR1 was highly expressed in dilated specimens and co-localized with CD68+ cells (Fig. 3C). Moreover, *OLR1* mRNA levels correlated positively with mRNA levels of inflammatory markers, in both dilated and non-dilated aortas (Fig. 3D). In addition, *OLR1* mRNA levels correlated positively with ascending aortic diameter (Fig. 3E).

**Figure 3.**
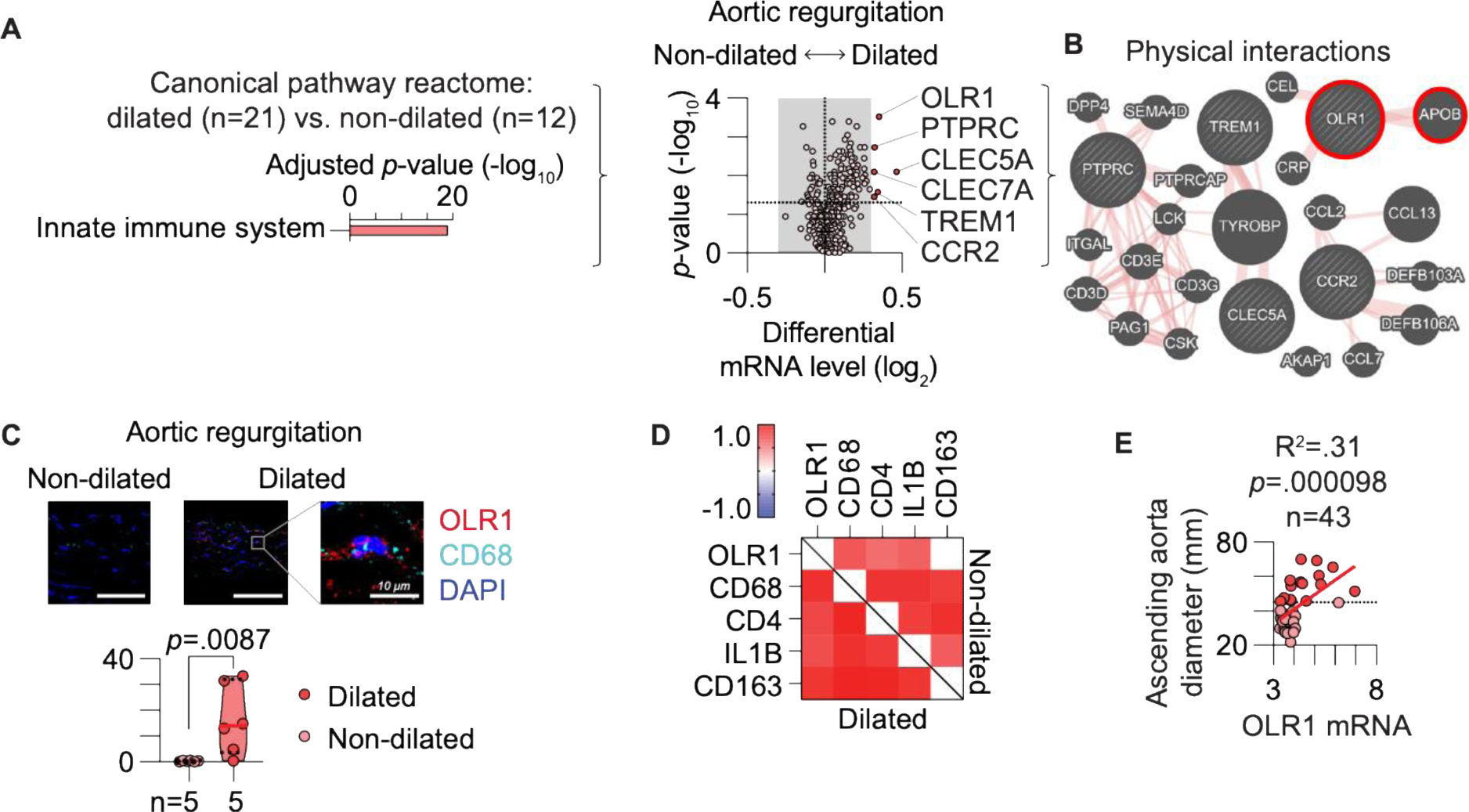
Immune cell OLR1 expression drives aneurysm progression through oxLDL- interactions **(A)** Pathway analysis of differentially expressed genes, dilated vs. non-dilated aortas of patients with aortic regurgitation. **(B)** Volcano plot of differentially expressed genes within the innate immune system term. **(C)** GeneMania physical interaction plot of top five differentially expressed genes. **(D)** Immunofluorescent double staining of CD68 and OLR1 in biopsies from dilated and non-dilated aortas of patients with aortic regurgitation, Mann- Whitney test. **(E)** Gene correlation matrix of Pearson R values, patients with aortic regurgitation. The upper right part of the matrix represents non-dilated aortas, and the lower left part representdilated aortas. **(F)** Linear regression analysis of ascending aortic diameter measured by perioperative echocardiography and OLR1 mRNA levels in patients with aortic regurgitation. Scale bar 100 µm in all micrographs. DAPI=4′,6-diamidino-2-phenylindole, LOX-1=Lectin-like oxidized lipoprotein receptor 1.

### Surgical aortic valve replacement for aortic regurgitation, but not aortic valve stenosis mitigates ascending aortic growth

As part of a follow-up study, in which ASAP-patients were re-called 10 years after ascending aortic and/or aortic valve surgery for evaluation of long-term aortic growth, we evaluated ascending aortic growth in relation to valve disease in patients undergoing isolated aortic valve replacement. Interestingly, aortic valve replacement for aortic regurgitation, but not aortic stenosis, mitigated further ascending aortic expansion (Fig. 4), with a mean growth of 0.86 mm vs. 1.67 mm for aortic regurgitation and aortic stenosis, respectively.

**Figure 4.**
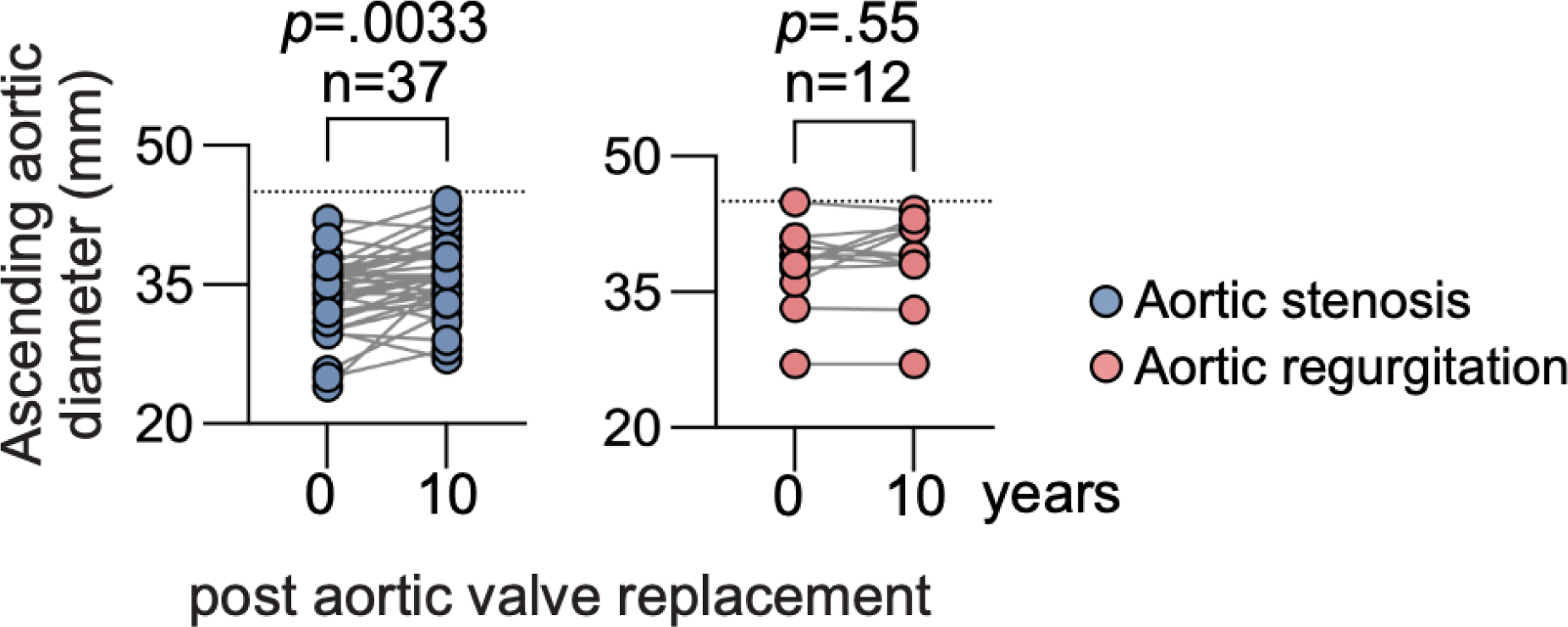
Surgical aortic valve replacement attenuates aortic growth in aortic regurgitation, not aortic stenosis Transthoracic aortic echocardiography follow-up data for ascending aorta diameter 10 years after aortic valve surgery and biopsy collection, Wilcoxon matched pair test.

## Discussion

Ascending aortic aneurysm is a silent, potentially fatal disease with no proven medical therapy. This is primarily due to a limited understanding of underlying pathological mechanisms and factors triggering the onset of disease. Here we show that the natural history of ascending aortic aneurysm formation in patients with tricuspid aortic valves involves subintimal infiltration of ApoB-carrying lipoproteins, which are retained and oxidized in the aortic wall. Up-regulation of oxidized low-density lipoprotein receptor 1 by infiltrating CD68+ cells further trigger an inflammatory cascade, most likely contributing to medial fibrosis and aneurysm formation. Importantly, the observed phenotype was only seen in patients with isolated aortic valve regurgitation, and not in patients with aortic stenosis who are mostly protected from aneurysm development. Surgical replacement of a regurgitant aortic valve mitigated further aortic expansion, whilst replacement of a stenotic valve was associated with a significant long-term aortic growth. Collectively this highlights oxLDL as a key player in the initiation of ascending aortic aneurysm formation and provides important insights towards improved disease prevention and management.

It has long been recognized that aortic regurgitation carries prognostic significance in conditions affecting the ascending aorta^24^, and histopathological studies have shown that aortic regurgitation is associated with medial degenerative insults already prior to aortic dilatation^14^. Here we further show that also genes encoding core proteins of LDL-binding proteoglycans^23^ are highly up-regulated in non-dilated aortas of patients with regurgitant valves, creating a favorable milieu for retention of infiltrating lipoproteins and thus, lipoprotein oxidation. Additionally, experimentally induced flow reversals, like those caused by a regurgitant aortic valve, has been reported to influence endothelial lipid biology^25^ and may contribute to the disease process.

The fact that expression of the atherogenic scavenger receptor Oxidized Low Density Lipoprotein Receptor 1 (*OLR1*) positively correlated with aortic diameter, and was significantly up-regulated in the aneurysmal aorta, also suggests a role of oxLDL in the progression of aneurysm formation. Indeed, *OLR1* mRNA levels correlated with the expression of inflammatory markers, in both dilated and non-dilated aortas, and its expression was co-localized with infiltrating CD68+ cells. Interestingly, innate immune cells are prone to greater inflammatory activation by oxLDL^26^, which in the context of AscAA, may exacerbate downstream immune cell-associated degeneration^6^. Furthermore, a subcluster of OLR1+ macrophages were recently demonstrated in dissecting aortic tissue, that promoted formation of neutrophil extracellular traps^27^. This indeed suggests a role for *OLR1* in the pathogenesis of ascending aortic aneurysm formation and warrants investigation as to whether OLR1 can be targeted pharmacologically as a preventive measure or diagnostic marker. Meanwhile, aortic valve replacement remains an effective, although invasive, therapy for patients with tricuspid aortic valves and aortic valve regurgitation, as indicated by the attenuation of long-term aortic growth following surgical valve replacement.

The present study also contests the controversial hypothesis of post-stenotic aortic dilatation^28^, where aortic stenosis *per se* is suggested to exacerbate ascending aortic growth. We could not see any infiltration of ApoB-containing lipoproteins, nor many CD68+ cells, into the subintima of patients with aortic stenosis, despite the fact that total cholesterol levels were higher in these patients compared with patients with regurgitant valves. It can be speculated that this may be due to high-velocity flow-jets imposed on the aortic wall as a result of aortic stenosis, limiting the exposure time of circulating lipoproteins to the endothelium and the transcytosis machinery^29,30^, and thus, hamper infiltration. However, interestingly, surgical replacement of a stenotic valve resulted in a significant ascending aortic growth over a period of 10 years. Although some aortic growth can occur without valvular abnormalities and with increasing age^31^, it is tempting to speculate that this may be a result of an adapted vascular smooth muscle cell phenotypic switch to compensate for the increased wall stress caused by the stenotic valve^13^, making the ascending aorta less resilient to pulsatile flow^32,33^. Indeed, an increased aortic stiffness has been observed in patients with aortic stenosis compared with patients with regurgitant valves following aortic valve replacement^34^.

## Limitations

The present report highlights novel results on the pathogenesis of degenerative AscAA but bases itself in a surgical cohort and therefore, limits the result to patients meeting the criteria for surgical intervention. Furthermore, longitudinal investigations do not consider changes in medical management of patients, and causality can be proven from the present study.

## Conclusion

Collectively, results from the present study suggest that oxidative modifications of lipoproteins and *OLR1*-expressing macrophages contribute to the early pathogenesis of ascending aortic aneurysm by propagating inflammation and matrix degeneration, ultimately leading to aortic dilatation. The causal relationships between regurgitant flow, lipid retention, and aortic dilatation, however, need to be confirmed.

## Ethics approval and consent to participate

The study was approved by the Human Research Ethics Committee in Stockholm (2006/784-31/1 and 2012/1633-31/4). Written informed consent was obtained from all patients following the declaration of Helsinki. The collection and scientific use of aortic tissues from organ donors was approved by the Human Research Ethics Committee in Stockholm (2017/2268-31).

## Acknowledgements

Not applicable.

## Data availability

The datasets used and/or analyzed in the current study are available from the corresponding author upon request.

